# Using a Simulation Centre to Evaluate the Effect of anArtificial Intelligence-Powered Clinical Decision Support System for Depression Treatment on the Physician-Patient Interaction

**DOI:** 10.1101/2020.03.20.20039255

**Authors:** David Benrimoh, Myriam Tanguay-Sela, Kelly Perlman, Sonia Israel, Joseph Mehltretter, Caitrin Armstrong, Robert Fratila, Sagar V. Parikh, Jordan F. Karp, Katherine Heller, Ipsit V. Vahia, Daniel M. Blumberger, Sherif Karama, Simone Vigod, Gail Myhr, Ruben Martins, Colleen Rollins, Christina Popescu, Eryn Lundrigan, Emily Snook, Marina Wakid, Jérôme Williams, Ghassen Soufi, Tamara Perez, Jingla-Fri Tunteng, Katherine Rosenfeld, Marc Miresco, Gustavo Turecki, Liliana Gomez Cardona, Outi Linnaranta, Howard C. Margolese

## Abstract

**Objective:** Aifred is an artificial intelligence (AI)-powered clinical decision support system (CDSS) for the treatment of major depression. Here, we explore use of a simulation centre environment in evaluating the usability of Aifred, particularly its impact on the physician-patient interaction.

**Methods:** Twenty psychiatry and family medicine attending staff and residents were recruited to complete a 2.5-hour study at a clinical interaction simulation centre with standardized patients. Each physician had the option of using the CDSS to inform their treatment choice in three 10-minute clinical scenarios with standardized patients portraying mild, moderate, and severe episodes of major depression. Feasibility and acceptability data were collected through self-report questionnaires, scenario observations, interviews, and standardized patient feedback.

**Results:** All twenty participants completed the study. Initial results indicate that the tool was acceptable to clinicians and feasible for use during clinical encounters. Clinicians indicated a willingness to use the tool in real clinical practice, a significant degree of trust in the AI’s predictions to assist with treatment selection, and reported that the tool helped increase patient understanding of and trust in treatment. The simulation environment allowed for the evaluation of the tool’s impact on the physician-patient interaction.

**Conclusions:** The simulation centre allowed for direct observations of clinician use and impact of the tool on the clinician-patient interaction prior to clinical studies. It may therefore offer a useful and important environment in the early testing of new technological tools. The present results will inform further tool development and clinician training materials.

## INTRODUCTION

Increasingly, new technologies which supplement clinical decision-making are being implemented to respond to the need to improve mental health treatment outcomes [1]. Some of these tools are designed to be used at the point of care, during sessions with patients, and may be expected to have some impact on physician-patient interactions. Impacts on these interactions may in turn affect the physician-patient relationship, one of the most critical aspects of psychiatric intervention [2]. It is challenging to test the effect of tools on these interactions and on clinical workflow; directly observing clinical interviews can be impractical or raise concerns about the validity of observations.

We assess the use of simulation to directly observe the impact of an artificial intelligence (AI)-powered decision support tool on simulated patient-clinician interactions. The objective was to determine if and how the use of the tool during a session impacted on the physician-patient interaction, as a prelude to longitudinal clinical studies assessing longer term effects on clinical workflow and the physician-patient relationship. Using simulation, clinician behaviour can be observed in a secure setting [3], and data collected from multiple viewpoints– that of the clinician, the standardized patient, and the observer. This triangulation process is a rigorous method for gathering high-quality data [4]. We discuss the challenges encountered and insights gained from our experience using simulation-based testing of new technology.

### AIFRED – CLINICAL DECISION SUPPORT SOFTWARE FOR DEPRESSION TREATMENT

We investigated the use of *Aifred*, a clinical decision support software (CDSS) which includes an operationalized version of the 2016 Canadian Network for Mood and Anxiety Treatments (CANMAT) guidelines for depression treatment [5] and provides AI decision support when treatments are chosen. This AI helps support clinicians by considering complex interactions between multiple patient variables to help personalize treatment in order to improve upon a trial-and-error treatment approach and reduce the number of failed treatment trials [6, 7]. It also tracks symptoms using standardized questionnaires such as the PHQ-9 [8]. Major depressive disorder (MDD) was chosen given its high prevalence [9, 10], status as the leading cause of disability globally [11], and poor remission rates following initial treatment [12].

The key innovation is the inclusion of an AI tool that provides clinicians with remission probabilities for different treatment options, based on a patient’s clinical and demographic profile. This AI is layered on top of the operationalized CANMAT guideline, providing remission probabilities for individual treatments at the point in the guideline when the first-line treatment is chosen. This AI tool is a deep-learning model trained and validated on baseline clinical and demographic data from 4,735 patients [6] from five major studies (STAR*D [12], CO-MED [13], EMBARC [14], REVAMP [15], and IRL-GREY [16]). It currently provides individualized remission probabilities for five commonly used first-line treatments (escitalopram, citalopram, bupropion, venlafaxine, and sertraline) as well as two combination treatments (bupropion plus escitalopram, and venlafaxine plus mirtazapine). In silico testing of this model demonstrated that it is likely capable of improving population remission rates (testing methods in [6]).

The tool is intended to be used during patient interviews, providing access to evidence-based decision support. Numerical remission probabilities are provided for those treatments on which the model is trained, but clinicians can choose from any of the treatments appearing in CANMAT. This simulation study sought to assess whether the tool, which should always be employed in the context of best clinical judgement and patient preference, could be feasibly used at the point of care while maintaining, or possibly enriching, the integrity of the physician-patient interaction.

## METHODS

The sample consisted of the intended end-users of the CDSS: psychiatry and family medicine attending staff and residents. Participants were recruited via email, social media, and announcements, and were compensated. The recruitment target was 25 participants. Recruitment started roughly three months before study start. The study was approved by the Research Ethics Board of the Douglas Mental Health University Institute. All participants provided informed consent.

The study was conducted at the Steinberg Centre for Simulation and Interactive Learning. Each participant was present at the simulation centre for one 2.5-hour session. The centre’s one-way mirror system allowed research assistants (RAs) to observe scenarios. The simulation centre has a roster of professional actors who play standardized patients (SPs). The ability of SPs to standardize their acting [17, 18], allows for multiple equivalent instances of the same clinical scenario to be run. RA’s wrote observations on data extraction forms created for the study.

We created three clinical situations, corresponding to a mild, moderate, and severe MDD. These situations were based on data from real patients drawn from the de-identified datasets on which the model was trained. “Jack” was a retired Caucasian male in his 80s suffering a mild depression marked by social withdrawal and sleep disturbance. He was experiencing some guilt about a previous divorce. “Emma” was a Caucasian professional female in her 40s suffering from moderate depression marked by agitation and guilt about poor performance at work and with respect to being emotionally available within her couple. “Sara” was an African-American female in her 50s who had lost her job due to a severe depression marked by psychomotor retardation and fatigue. She was prompted to come in to see the doctor by her friends in the building where she lives. The CDSS provided different remission probabilities per treatment for each patient.

Participants arrived in groups of up to six, and were given an introductory session that covered the current state of depression treatment, the rationale for the development of an AI-powered tool, current results of the AI model, and an introduction to the user interface of the tool. They were told that the SPs were playing patients who had used the tool to fill out questionnaires in the “waiting room” but had limited knowledge of the tool.

Participants were paired with an RA, who guided them through a 10-minute training session with the CDSS on a laptop. Participants then filled out a questionnaire recording their initial impressions of the tool. Each participant then interacted with all 3 standardized patients in a random order in three 10-minute clinical scenarios. During scenarios participants were free to interact with a laptop computer running the CDSS. The laptop was angled at 45 degrees towards the participant but could be freely moved to face the SP. The CDSS had access to questionnaire results as well as the treatment algorithm with its integrated AI tool. Participants were warned that as scenarios were only 10 minutes long, they should consider starting to use the CDSS roughly halfway through, however they were also told that they had the freedom to use or ignore the CDSS as they saw fit. After each scenario, participants filled out a questionnaire about their experience using the CDSS.

After the scenarios, there was a 10-minute structured interview with an RA in which participants were able to elaborate further on their experience. They were then asked to complete an anonymous “exit” questionnaire summarizing their experience using the tool and their opinion of its impact on the physician-patient interaction. The last step was a 10-question surprise quiz on the CANMAT 2016 Guidelines for Depression Treatment, intended to establish participant knowledge of guidelines. After each testing day, an unstructured debriefing session was held with all SPs. While SP feedback is often not standardized, SPs have been shown to effectively assess clinical skills [17, 19, 20], which motivated us to consider SP feedback when assessing the impact of the tool on the clinician-patient interaction.

## RESULTS

Results are derived from the registration and exit questionnaires unless otherwise noted, and comment on participant satisfaction, knowledge and skills gained, and potential impact on clinical practice. Note that these are initial selected results meant to illustrate the utility of the simulation centre; full study results will be reported separately.

Twenty participants completed the study. Participants were nearly evenly split between psychiatry (n = 11) and family medicine (n = 9), with a wide age range (24-67, mean age 39.5) and practice experience (6 residents and the following breakdown in experience for attending staff: 0-5 years: 4, 6-10 years: 2, 11-15 years: 2, 16-20 years: 4, 21+ years: 2). The sample included participants practicing in hospital and community settings.

With respect to participant satisfaction and impact on the physician-patient interaction, 70% of participants felt that the AI model assisted them in helping their patients better understand treatment (scoring ≥ 4 on a 1-5 scale, with higher values representing greater confidence). Sixty-five percent felt it helped improve patient trust in the treatment (scoring ≥ 4 on a 1-5 scale). Fifty percent felt that the application provided them with richer information to discuss with their patients (≥ 4 on a 1-5 scale). Forty-five percent of participants reported that using the application made the interaction with patients feel less personal or that it interfered with their interview (≥4 on a 1-5 scale). Seventy percent felt the remission probabilities provided by the model were reasonable overall.

In terms of potential impact on clinical practice, 50% of participants thought they would use the CDSS for all of their patients with MDD, with an additional 40%, therefore 90% overall, stating they would use it for more complex or treatment-resistant patients. Sixty percent of participants trusted that the AI could help them choose treatments (≥4 on a scale 1 to 5). Eighty percent of participants felt that the information on the treatment selection page in the application (which indicated CANMAT-recommended treatments, their usual doses, as well as the AI predictions) contained information that was clinically useful (≥4 on a scale 1 to 5). This suggests that the information contained in the tool could augment clinician knowledge during their interactions with patients. See Table 1 for a summary of results.

**Table 1.**
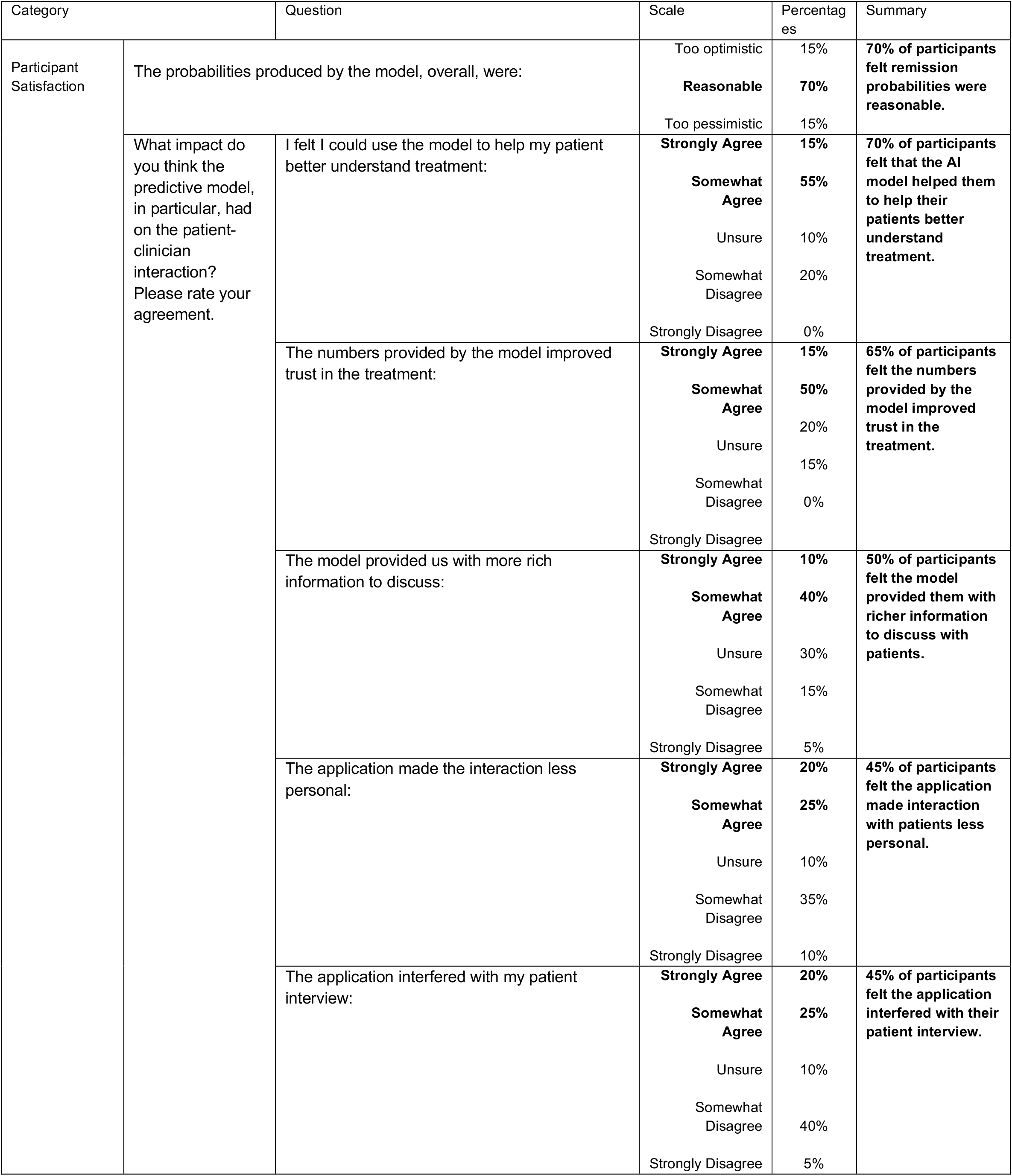
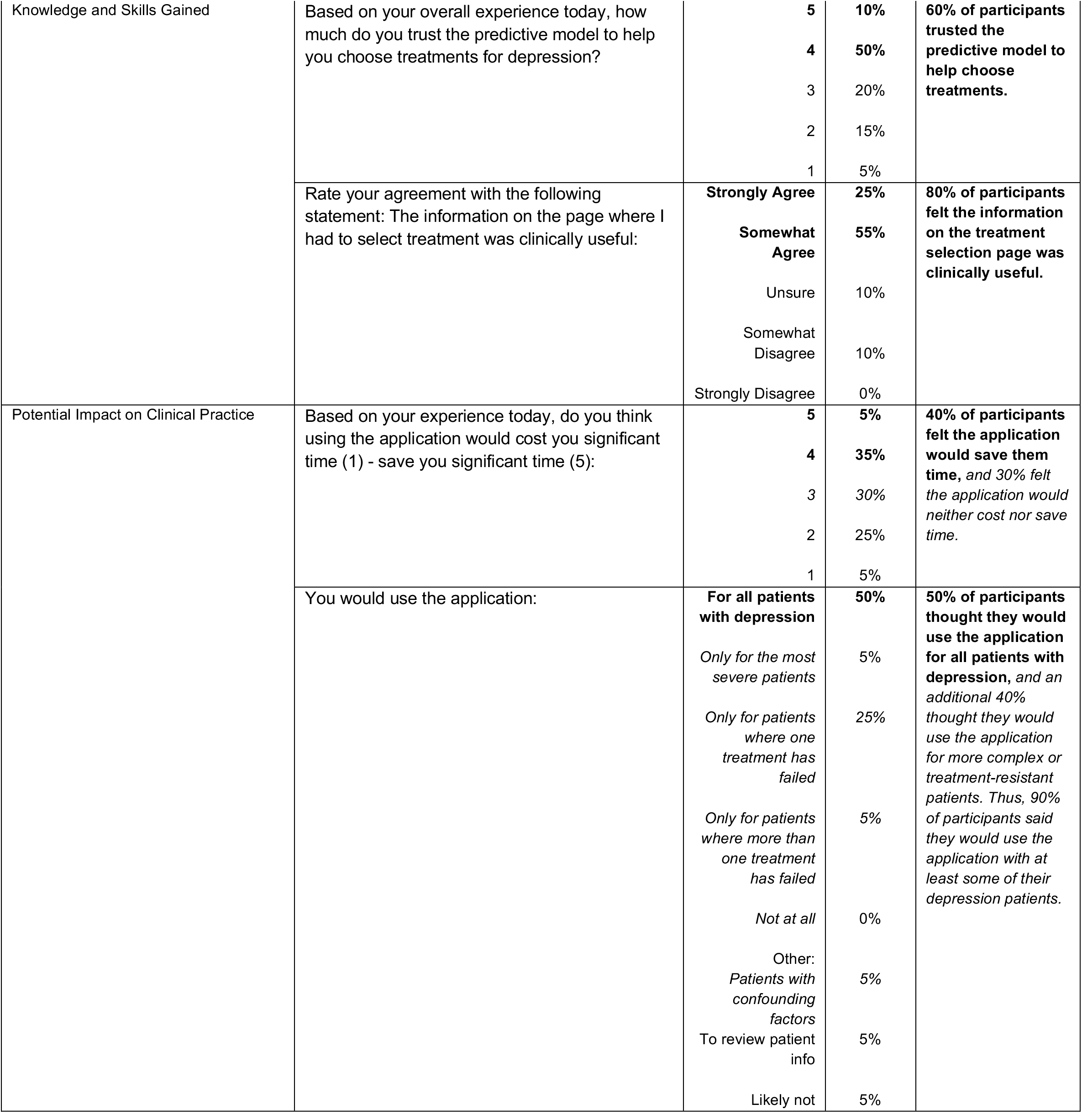
Study results by category

| Category | Question |  | Scale | Percentages | Summary |
| --- | --- | --- | --- | --- | --- |
| Participant Satisfaction | The probabilities produced by the model, overall, were: |  | Too optimistic | 15% | **70% of participants felt remission probabilities were reasonable.** |
|  |  |  | **Reasonable** | **70%** |  |
|  |  |  | Too pessimistic | 15% |  |
|  | What impact do you think the predictive model, in particular, had on the patient-clinician interaction? Please rate your agreement. | I felt I could use the model to help my patient better understand treatment: | **Strongly Agree** | **15%** | **70% of participants felt that the AI model helped them to help their patients better understand treatment.** |
|  |  |  | **Somewhat Agree** | **55%** |  |
|  |  |  | Unsure | 10% |  |
|  |  |  | Somewhat Disagree | 20% |  |
|  |  |  | Strongly Disagree | 0% |  |
|  |  | The numbers provided by the model improved trust in the treatment: | **Strongly Agree** | **15%** | **65% of participants felt the numbers provided by the model improved trust in the treatment.** |
|  |  |  | **Somewhat Agree** | **50%** |  |
|  |  |  | Unsure | 20% |  |
|  |  |  | Somewhat Disagree | 15% |  |
|  |  |  | Strongly Disagree | 0% |  |
|  |  | The model provided us with more rich information to discuss: | **Strongly Agree** | **10%** | **50% of participants felt the model provided them with richer information to discuss with patients.** |
|  |  |  | **Somewhat Agree** | **40%** |  |
|  |  |  | Unsure | 30% |  |
|  |  |  | Somewhat Disagree | 15% |  |
|  |  |  | Strongly Disagree | 5% |  |
|  |  | The application made the interaction less personal: | **Strongly Agree** | **20%** | **45% of participants felt the application made interaction with patients less personal.** |
|  |  |  | **Somewhat Agree** | **25%** |  |
|  |  |  | Unsure | 10% |  |
|  |  |  | Somewhat Disagree | 35% |  |
|  |  |  | Strongly Disagree | 10% |  |
|  |  | The application interfered with my patient interview: | **Strongly Agree** | **20%** | **45% of participants felt the application interfered with their patient interview.** |
|  |  |  | **Somewhat Agree** | **25%** |  |
|  |  |  | Unsure | 10% |  |
|  |  |  | Somewhat Disagree | 40% |  |
|  |  |  | Strongly Disagree | 5% |  |

|  |  |  |  |  |
| --- | --- | --- | --- | --- |
| Knowledge and Skills Gained | Based on your overall experience today, how much do you trust the predictive model to help you choose treatments for depression? | 5<br>4<br>3<br>2<br>1 | 10%<br>50%<br>20%<br>15%<br>5% | 60% of participants trusted the predictive model to help choose treatments. |
|  | Rate your agreement with the following statement: The information on the page where I had to select treatment was clinically useful: | Strongly Agree<br>Somewhat Agree<br>Unsure<br>Somewhat Disagree<br>Strongly Disagree | 25%<br>55%<br>10%<br>10%<br>0% | 80% of participants felt the information on the treatment selection page was clinically useful. |
| Potential Impact on Clinical Practice | Based on your experience today, do you think using the application would cost you significant time (1) - save you significant time (5): | 5<br>4<br>3<br>2<br>1 | 5%<br>35%<br>30%<br>25%<br>5% | 40% of participants felt the application would save them time, and 30% felt the application would neither cost nor save time. |
|  | You would use the application: | For all patients with depression<br>Only for the most severe patients<br>Only for patients where one treatment has failed<br>Only for patients where more than one treatment has failed<br>Not at all<br>Other:<br>Patients with confounding factors<br>To review patient info<br>Likely not | 50%<br>5%<br>25%<br>5%<br>0%<br>5%<br>5%<br>5% | 50% of participants thought they would use the application for all patients with depression, and an additional 40% thought they would use the application for more complex or treatment-resistant patients. Thus, 90% of participants said they would use the application with at least some of their depression patients. |

Prior to the simulation, 75% of participants reported that they would realistically use the application in clinic for five minutes or less during a session. Forty percent of participants reported that the application would save them time (≥4 on a scale 1 to 5), and 30% felt the application would neither save nor cost them time (3 on a scale 1 to 5), indicating potential feasibility in a real, busy clinical environment. This was corroborated by the fact that, in the majority of scenarios, the participants were able to successfully navigate through the application within the short time provided. In a questionnaire administered right after each clinical scenario, 61.7% of participants reported that using the application “took some adjustment, but […] worked well”. SPs provided valuable feedback, such as noting that some participants turned the computer screen towards them during the session, “inviting them in” to engage with the tool. This seemed to be linked to acceptability of the tool’s presence on the part of the SPs. They also commented on the importance of the clinician’s manner and rapport building skills, such as warmth and ability to engage them in their care.

## DISCUSSION

We will now reflect on the use of simulation for testing the effect of new technologies on the physician-patient interaction. Our initial results demonstrate that a majority of clinicians were satisfied with the use of the CDSS. At the end of the simulation, most clinicians could see themselves using the tool for at least a subset of their patients with depression, suggesting the feasibility of using the tool to achieve real-world impact. No major threats to the quality of the physician-patient interaction were identified, and we illustrated several ways in which the tool might enhance the interaction, as well as tools clinicians can use to better integrate the CDSS into a session.

Our sample of 20 participants was diverse with respect to career stage and practice environment, which increases confidence in the generalizability of our results. The sample size reflects recruitment feasibility. The largest barrier to recruitment was clinical duties - and for residents, concerns about not being released to participate. Being able to offer more testing days, as well as departmental approval for residents’ participation, may have increased recruitment. A challenge with simulation is that running participants in groups on predefined days is necessary given the need to ensure room and SP availability.

Using simulation-based testing allowed us to observe interactions that would not have been easily accessible in other settings. As noted, some participants tended to turn the laptop towards their SP. SPs referred to this as participants “inviting them in;” this behaviour seemed to be important in determining SP experience of the tool. SP feedback and our observations of sessions revealed that traditional aspects of the physician-patient interaction - such as clinician warmth, body language and ability to engage the patient - were also important in determining the SP experience, suggesting that the impact of a new technology may depend on clinicians’ baseline ability to build rapport with their patients. This merits further investigation in a clinical environment. Self-report from clinicians also revealed important impacts of the CDSS on the physician-patient interaction, such as the perceived utility of the tool in helping them better explain and increase trust in treatment. This interplay of observations of clinician behaviour, clinician self-report and SP experiences provided fundamentally different information than would have been obtained through clinician self-report alone. These observations will influence clinician training provided in future clinical studies, resulting in more focus on how clinicians can engage the patient with the tool in-session and use it to provide more information and enhance patient trust.

External validity is a concern when using simulation-based testing [3]. For example, several participants noted in written comments and during interviews that the 10-minute training session was insufficient and that they would likely have become more comfortable with the CDSS with more time. However, external validity may depend on research aims [3]. In our case, the aim was to see if the application was intuitive to use with minimal training, and, as noted, the majority of participants felt the tool took some adjustment but worked well. Similarly, the 10-minute clinical session length was felt by multiple participants to be too short. We initially hypothesized that most clinicians would want a tool that they could use in five minutes and this was supported by the finding that, at baseline, 75% of participants could see themselves using the tool for 5 minutes or less. Having short sessions, in which most participants used the tool in the latter half of the session, allowed us to determine that it is possible to use the tool in a meaningful way within this time constraint. As such our research aims were well suited to simulation work.

The use of the simulation environment - and crucially of SPs - to test the impact of technology on the physician-patient interaction is both practically useful and important as it allows direct observation of clinician interaction with a new tool prior to patient studies. This method provides multiple points of observation, allowing for an informative and multi-faceted dataset which can inform the development of tools and training materials. Evaluating the ease with which new technology is used and integrated into clinical practice is a key step in the proper development and implementation of novel clinical tools, and is a useful prelude to more longitudinal studies on the impact of these tools on the clinician-patient relationship.

## Data Availability

The data is not currently publicly available.

## Acknowledgements

We would like to acknowledge the Steinberg Centre for Simulation and Interactive Learning for the helpfulness of their staff in assisting with the execution of this study, as well as the standardized patients (SPs) who participated in the study for their excellence and the quality of their feedback.

